# Reach and uptake of facility- and home-based hepatitis C self-testing in urban slums in Haridwar, India; A Comparative Implementation Study

**DOI:** 10.64898/2026.09.27.26364104

**Authors:** Ajeet Singh Bhadoria, Amrita Mehndiratta, Yasmin Dunkley, Kathirvel Selv, Rajesh Somvanshi, Muhammad Shahid Jamil, Niklas Luhmann, Cheryl Johnson, Karin Hatzold

## Abstract

Routine facility-based testing for hepatitis C virus (HCV) remains underutilised in India despite national screening policy. WHO has recommended HCV self-testing (HCVST) since 2021 to increase access to testing services. This study evaluated the HCVST by assessing its reach and uptake in urban-slum communities within Haridwar, Uttarakhand, India.

The implementation study (CTRI/2023/07/054590) took place between 12^th^ July 2023 and 31^st^ August 2024. Three Primary Healthcare Centre (PHC) catchment areas were assigned to deliver either (1) facility-based HCVST, (2) home-based HCVST, or (3) standard of care (SoC). Facility-based delivery involved trained users offering kits to patients visiting the PHC (Site 1: Jwalapur 1), whereas home-based delivery (Site 2: Jwalapur 2) involved trained users and ASHAs approaching community households selected via a KISH grid. Primary outcomes were OPD-footfall standardized operational reach; the proportion offered testing out of the PHC catchement areas footfall, and uptake (acceptance among those offered). Secondary analysis explored demographic and behavioural predictors of those that refused to test.

Of 7,091 outpatient department (OPD) footfall, 724 (10.2%) individuals were offered HCVST at intervention sites, of which 465 (64.2%) agreed to self-test. Operational reach was 17.8% (393/2,212; 95% CI: 16.2–19.5%) through the facility-based intervention and 14.4% (331/2,296; 95% CI: 13.0–15.9%) in the home-based intervention.HCVST uptake was higher through the home-based model (71.9%, 238/331, 95% CI: 66.7–76.7%) than in the facility (57.8%, 227/393, 95% CI: 52.7–62.7%). At the SoC site, only 11 individuals received standard of care HCV testing. The study HCV positivity was 1.1% (5/465; 95% CI: 0.35%– 2.49%): with four testing positive at the facility site (4/227), one at the community site (1/238), and none at the SoC site (0/11). Men were more likely to refuse HCVST than women (OR 0.54, 95% CI 0.37–0.78, p=0.001).Participants in the facility-based model had 1.87 times higher odds of refusal compared to those in the home-based delivery model.

There is minimal routine HCV testing in the OPD setting of Urban PHCs despite national screening policy. HCVST improved testing coverage and uptake in urban slums compared to SoC. However, HCV positivity was low overall, suggesting that untargeted screening – whether facility-based or home-based - may identify few infections without additional strategies to reach higher-risk populations.

## Introduction

The World Health Organization (WHO) has set ambitious targets to eliminate hepatitis C virus (HCV) as a public health threat by 2030, aiming to diagnose 90% of infections and treat 80% of those diagnosed (1). Aligning with these goals, India launched the National Viral Hepatitis Control Programme (NVHCP) in 2018. At the Primary Health Care (PHC) level, the focus is on prevention through health education, behavioral change communication regarding safe injection and blood practices, and free opportunistic screening using rapid diagnostic tests (RDTs) (2). However, a stark divide exists between official policy and realities on the ground. While the national strategy intends for opportunistic screening to be offered to symptomatic individuals or those with risk factors, implementation is fragmented; in practice, screening is largely limited to pregnant women attending antenatal care (ANC), leaving the broader outpatient department (OPD) population largely unreached (3).

Governance and policy constraints, such as variable state prioritization, insufficient accountability, and a lack of robust surveillance data, further limit the program’s ability to reach target populations (4,5).

National estimates suggest adult HCV prevalence in India ranges from 0.32% to 3.23% (2). According to the WHO Global Hepatitis Report 2024, an estimated 5.5 million people in India are living with HCV, representing 11.2% of the total global burden. This disproportionate contribution emphasizes the urgent need for expanded, proactive screening strategies (6). Localized data from Uttarakhand also highlights regional burden; a 2013 study in a tertiary care setting reported an HCV seroprevalence of 1.8% (9/495), closely approaching the WHO-defined threshold of >2% for universal general population screening (7,8). Haridwar, a rapidly urbanizing district in Uttarakhand, exemplifies these challenges.

The district contains 41 urban slums accommodating approximately 25% of the city’s population (9). While geographically proximate to health centers, urban slum residents remain functionally excluded due to structural barriers, including the prohibitive cost of losing a day’s wage and the profound social stigma associated with a positive diagnosis (2,10). Furthermore, many urban slum residents rely on informal healthcare providers where unsafe practices, such as needle/syringe reuse, significantly elevate transmission risks (11,12). Migration to urban slums may also increase vulnerability to HCV infection; occupational exposures among rural-to-urban migrants in other contexts have been associated with HCV prevalence rates up to 3.8 times higher than the general population suggesting the need for tailored diagnostic strategies (13).

Reaching these populations requires a shift from passive, facility-centric models to active, person-centric strategies. In 2021, the WHO recommended HCV self-testing (HCVST) as an additional strategy to increase testing uptake (10). HCVST can be delivered through multiple service delivery pathways, including distribution within health facilities or through community-based approaches that bring testing directly to households. These approaches may influence both the number of individuals reached and the likelihood that those offered testing will accept it. This study evaluates two HCVST delivery model; facility-based self-testing and home-based distribution delivered through community peers called Accredited Social Health Activists (ASHAs) to facilitate household and point-of-care distribution to examine how different service delivery approaches influence testing reach and uptake in underserved urban slum communities.

## Methods

### Study design

Cross-sectional analysis of an implementation study was conducted. Data is reported in accordance with STROBE guidelines (14,15).

### Setting

Between 12^th^ July 2023 and 31^st^ August 2024, three PHCs were selected randomly in the Haridwar district of Northern India: Kankhal, Jwalapur 1 and Jwalapur 2. Baseline site assessments confirmed that these three facilities were sufficiently comparable regarding human resources, staff training and average patient footfall. Furthermore, all three sites demonstrated similar levels of population coverage and baseline utilization of hepatitis C screening kits, diagnostics, and referral to higher facilities for treatment (as provided in Supplement 1, **S1**). This equivalence ensured that the extent of service delivery under the National Viral Hepatitis Control Programme (NVHCP) was uniform across sites prior to the introduction of the study interventions.

### Intervention and implementation approach

#### Intervention

The First Response HCV Card Test (Premier Medical Corporation Pvt Ltd., Gujarat, India) was used for self-testing. Instructions for Use pamphlets were provided in English and Hindi. Self-testing was supervised; in the facility-based model by study staff, and in the home-based model by the Accredited Social Health Activists (ASHAs). Supervision was a study requirement because these kits had not yet obtained WHO pre-qualification for self-testing use. The entire sequence, spanning from the recruitment of the participant to the final interpretation of the result, typically required between 15 and 20 minutes to complete. Venous blood collection were collected from all 465 participants who completed HCVST for independent verification using laboratory-based anti-HCV ELISA. Participants with reactive HCVST results additionally underwent HCV RNA testing to determine active infection.

Those with detectable HCV RNA were referred to the nearest government treatment facility for clinical evaluation and initiation of direct-acting antiviral therapy and were followed up by telephone through assessment of sustained virological response 12 weeks after treatment completion.

#### Study Eligibility

Any adult (≥18 years old) resident of the urban slums for at least six months prior to the intervention within a selected PHC catchment area was eligible to access testing. Those who had a known HCV diagnosis (on treatment or cured) were excluded from the study.

#### Implementation

Each PHC was randomly allocated to deliver a different delivery model.

**Kankhal** was designated as SoC, continuing its provision of routine NVHCP activities; these comprised provision of rapid diagnostic kits for HCV screening, health awareness sessions conducted during special health days or Patient Welfare Committees (Rogi Kalyan Samiti) - a health facility level, community-based hospital management meeting held fortnightly (19), as well as during Village Health and Sanitation Days/Nutrition Weeks.

**Jwalapur 1** was allocated the facility-based HCVST delivery model. In addition to routine NVHCP activities, HCVST kits were made available. Over seven months of implementation (July 2023 - January 2024), trained study staff recruited participants and provided them with

HCVST kits, standard instructions and educational pamphlets in Hindi language. Participants then performed the self-test under active supervision by the study staff.

**Jwalapur 2** was allocated home-based HCVST delivery model using a trusted community peer. ASHAs recruited participants during routine household survey visits. Normally, an ASHA’s role is limited to providing health education and generating awareness regarding the availability of free testing and treatment services under NVHCP at primary healthcare centres. Whereas in this intervention model, ASHAs supervised self-testing. Eligible individuals within each household were identified using the Kish Grid method (16).

Table 1 summarises the implementation procedures across the three study sites.

**Table 1.** Implementation characteristics of the study sites.

| **Implementation feature** | **Facility-based HCVST** | **Home-based HCVST** | **Standard of care** |
| --- | --- | --- | --- |
| Study site | Urban PHC Jwalapur 1 | Urban PHC Jwalapur 2 catchment area | Urban PHC Kankhal |
| Delivery setting | Within the Urban PHC outpatient department | Participants' households within the UPHC Jwalapur 2 catchment area | Routine Urban PHC outpatient services |
| Target Population | OPD attendees aged $\geq 18$ years met the study eligibility criteria | Community residents aged $\geq 18$ years met the study eligibility criteria | OPD attendees receiving routine care under the NVHCP; No systematic study-specific eligibility screening |
| Recruitment approach | Consecutive approach to patients attending the OPD | Households and eligible adults selected using the Kish grid | Routine NVHCP service delivery; no active HCVST recruitment; Testing was based on routine clinical assessment and provider judgement and, where applicable, patient request |
| Personnel involved | Trained HCVST provider and laboratory technician, with support from the medical officer | Trained HCVST provider and laboratory technician, supported by ASHAs | CHOs, ANMs and ASHAs under routine programme services |
| Service promotion | Individual counselling and pamphlet distribution among OPD attendees | Individual counselling during ASHA outreach and household visits | Routine NVHCP awareness and behaviour-change communication<br>No dedicated HCVST personnel or study-supported active recruitment; testing continued through routine programme staffing and resources |
| Testing procedure | Supervised self-testing and interpretation at the facility; | Supervised self-testing and interpretation at home; result | Provider-administered facility-based HCV screening using HCV Rapid Diagnostic Tests (RDTs); no self- |
|  | result verified by the trained provider | verified by the trained provider | testing |
| Confirmatory Testing | Venous blood was collected from all participants who completed HCVST for Laboratory based anti-HCV ELISA. Reactive results underwent HCV RNA testing to determine active infection |  | Individuals with reactive routine HCV rapid diagnostic test results were managed according to the routine NVHCP confirmatory-testing pathway. |
| Referral and Linkage Follow-up | Participants with confirmed active HCV infection were linked to the nearest government treatment facility and followed up by telephone through SVR12 assessment. |  | Routine NVHCP referral and linkage-to-care pathways. |

A more detailed description of the implementation activities is reported in accordance with the Template for intervention description and replication (TIDieR) checklist (S2) (17).

### Sample Size and Sampling

No published data from India were available to inform expected HCVST uptake, therefore a pragmatic sample size was calculated to detect a 20-percentage-point absolute difference in uptake between the two HCVST delivery models. Assuming uptake proportions of 60% and 40%, a two-sided significance level of 5%, 80% power, and equal allocation, 102 individuals were required per group. After applying a design effect of 2 to account for clustering and allowing 10% for incomplete data, the minimum required sample was 227 individuals per group. For field implementation, an operational target of approximately 230 completed HCVSTs per delivery model was set. HCVST was offered until this completion target of 460 was reached. Consequently, 393 individuals were offered HCVST in the facility-based model and 331 in the home-based model. All 724 individuals offered HCVST contributed to the uptake analysis, of whom 465 completed HCVST: 227 in the facility-based model and 238 in the home-based model.

### Outcome and exposure variables

Across the HCVST intervention sites, the primary outcomes were operational reach and uptake. To provide a common descriptive benchmark for implementation volume across the two HCVST delivery models, operational reach was measured using OPD-footfall-standardized delivery coverage. This was defined as the number of individuals offered HCVST at each intervention site divided by the total number of people who attended the outpatient department (OPD) recorded at the corresponding Urban PHC over the same time-period. For the facility-based model, the denominator represented the OPD footfall at Urban PHC Jwalapur from which individuals were offered HCVST. For the home-based model, OPD footfall at the Urban PHC serving the intervention catchment area, Jwalapur 2, was used as a standardized reference measure of local facility utilisation. This denominator served as a proxy for catchment-level service volume and did not represent the number of community contacts approached, screened for eligibility for HCVST. Therefore, operational reach in the home-based model was interpreted as the scale of community HCVST delivery relative to routine facility utilisation and not as population-level community reach.

Uptake was defined as the proportion of individuals who accepted HCVST among those who were offered the test during the same period.

Exposure data was extracted from a study tool administered by trained research staff prior to the intervention. Exposure variables included in this analysis were gender (male/ female), age group (18-30, 31-45, 46-60 and >60) and educational level (primary school, middle school, high school, intermediate school/ diploma and higher education), both grouped to provide statistical stability for regression analysis. Socioeconomic Status (SES) was constructed through the modified B.G. prasad SES scale 2024 which classifies SES based on per capita monthly income though five classes, where class I is the highest SES status to class V the lowest (18). Marital status (Married or living with a partner, widowed, divorced or separated, or never married) was grouped in line with census data.

To document reasons for refusal, participants were provided with a description of the study and the HCV self-testing procedure, followed by a request for consent. If they declined, an open-ended question in Hindi (e.g., *“Could you share why you do not wish to participate?”*) was asked to capture their rationale. Responses were later thematically analysed and categorized into distinct refusal reasons.

### Statistical Analysis

We graphed flow diagrams depicting participant flow throughout the study by site. We described absolute numbers offered self-tests and numbers taking up self-tests, and respective proportions with binomial exact confidence intervals, and compared numbers between ST sites. We summarised participant characteristics as categorical using proportions and compared variables between sites using chi square tests.

To explore factors associated with testing refusal, we compared the demographic characteristics of individuals who declined HCV self-testing with those who accepted testing using chi-square tests for categorical variables and t-tests for continuous variables. We then fitted a binary logistic regression model to examine predictors of testing refusal among individuals who were offered HCVST. The outcome variable was refusal of HCVST, age, gender, and study site/mode of HCVST delivery model were included as explanatory variables. Odds ratios (ORs) with 95% confidence intervals were estimated.

We calculated the overall case detection proportion, and stratified this between HCVST-sites and SoC-sites using chi square tests. We also described linkage to care and achievement of sustained virologic response (SVR), i.e., the propoprtion of reactive results who were successfully linked to confirmatory testing, initiated treatment, and achieved cured. All statistical analyses were performed using Stata version 18 (StataCorp LLC, College Station, TX, USA).

### Ethical Considerations

Informed written consent was obtained from all participants. Ethical approval was obtained from the Institutional Ethics Committee of AIIMS Rishikesh reference no. 78/IEC/EM/2023 prior to study initiation.

## Results

### Study throughput

During the study period, a total of 7,091 individuals attended the OPDs of the three selected PHCs in Haridwar district. At the site allocated facility-based HCVST (Jwalapur 1), the total OPD footfall was 2,212. At the site allocated the home-based HCVST delivery (Jwalapur 2), the OPD footfall was 2,296. At the Standard of Care site (Kankhal), the OPD footfall was 2,583 (**Fig 1**).

**Figure 1.**
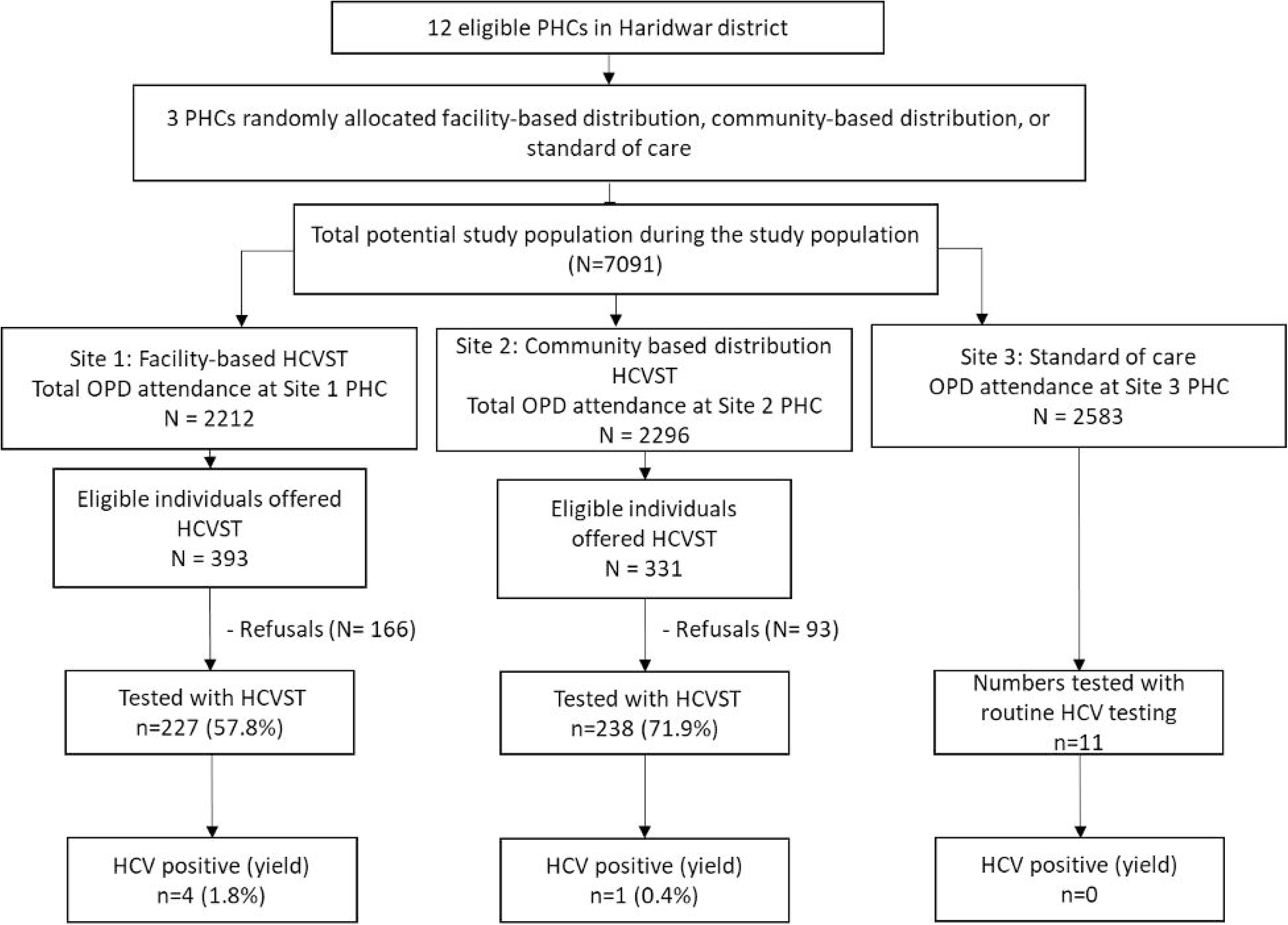
Study throughput

### Reach and Uptake

Across the two HCVST intervention sites, a total of 724 people were offered HCVST as part of this study out of a total OPD footfall of 4508 representing a total operational reach of 16.1% (724/4508, 95% CI: 15.0 % – 17.2 %) of the OPD population. By site, the facility-based HCVST reached a greater proportion of the OPD footfall (17.8%, 393/2,212; 95% CI: 16.2 – 19.5%) compared to the home-based HCVST delivery site (14.4%, 331/2,296; 95% CI: 13.0–15.9%).

Among those offered self-testing, a total of 465 agreed to use the self-test, which translates to an uptake of around two-thirds (64.2%, 465/724, 95% CI: 60.6 % – 67.7%). While a slightly higher proportion of individuals were reached with the HCVST offer through facility-based HCVST, the uptake was greater among those offered the test through home-based HCVST (71.9%, 238/331; 95% CI: 66.7–76.7%) compared to facility-based HCVST testing (57.8%, 227/393; 95% CI: 52.7–62.7).

At the SoC site, only 11 individuals underwent HCV rapid diagnostic testing out of 2583 OPD population over the same time period.

### Participant demographics

There was roughly equal gender breakdown amongst the HCVST participants; just under half participants were males (46.5%, 216/465), with no significant difference in distribution between the facility-based (45.4%, 103/227) and home-based models (47.5%, 113/238; p = 0.717). The participants in the facility-based model were significantly younger than those in the home-based model (mean age 36.1 ± 13.7 vs. 40.4 ± 16.4 years; p = 0.002). Accordingly, the categorical age distribution also differed significantly between the two delivery models (p = 0.004); the 18–30 age group comprised a larger proportion of the facility-based model (45.4%, 103/227 vs. 34.0%, 81/238), whereas the home-based model reached a larger proportion of older adults >60 years (16.4%, 39/238 vs. 7.0%, 16/227).

Overall, a quarter of participants had primary education level or lower (25.4%, 118/465), showing no significant difference by site (24.2%, 55/227 at facility-based site vs. 26.5%, 63/238 at home-based site; p = 0.214). Just under half (42.2%, 196/465) of the participants belonged to Class IV (the second lowest SES); however, the overall socioeconomic status distribution differed significantly between the two sites (p = 0.0301). More than a third of participants had no reported history of risk exposure (35.5%, 165/465), with no significant difference by site (33.5%, 76/227 at facility vs. 37.4%, 89/238 at home; p = 0.4325), though a history of a surgical procedure was reported significantly more frequently by participants in the home-based model (32.8%, 78/238) than by those in to the facility-based model (15.9%, 36/227; p < 0.001) (**Table 2**).

**Table 2.**
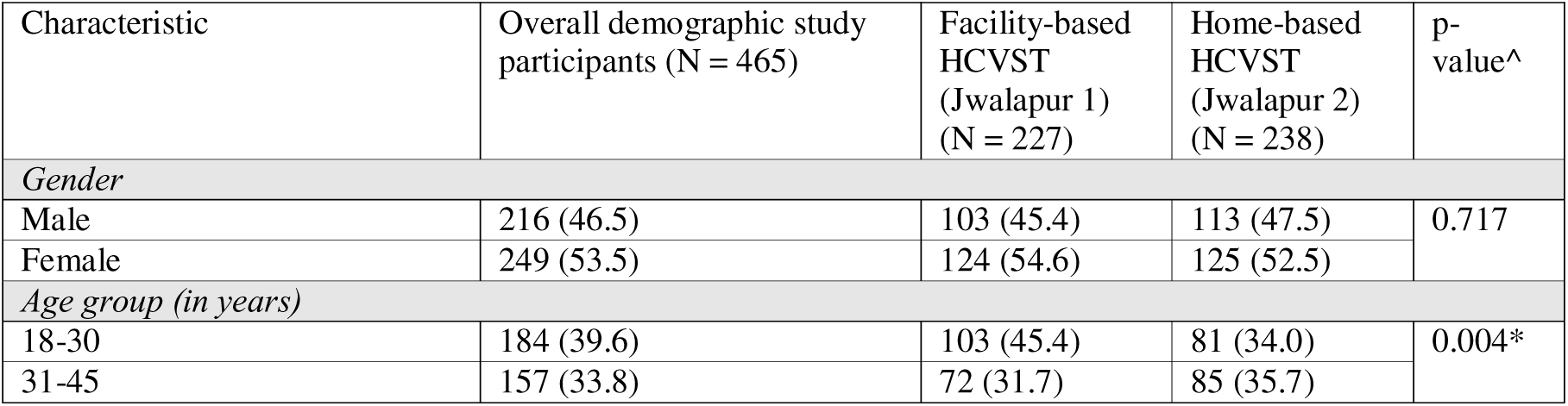
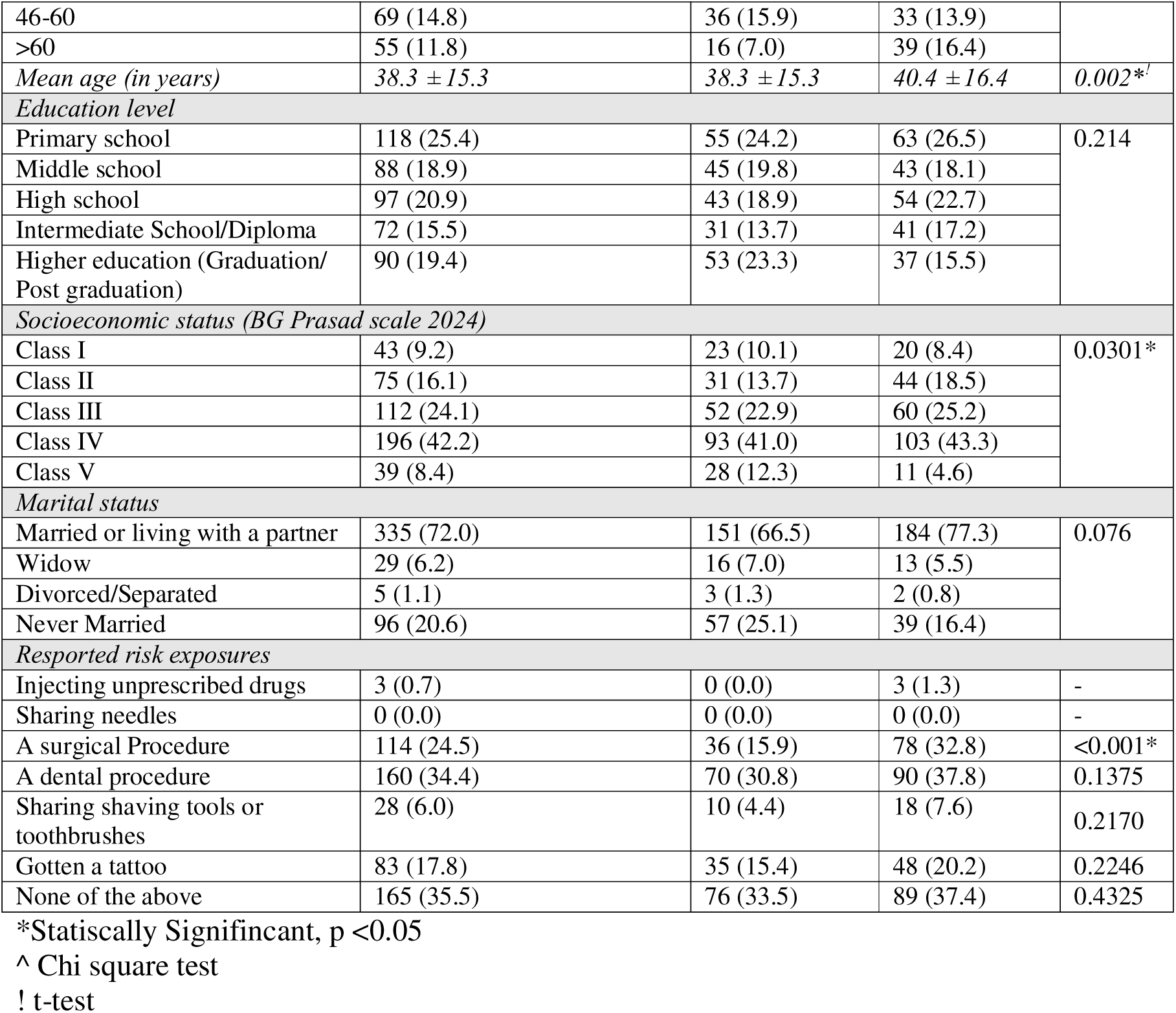
Demographic Characteristics of the HCVST study participants.

### Factors associated with testing refusal

To explore factors associated with testing refusal, we compared the demographics of individuals who declined HCV self-testing with those who agreed to test. Of the 724 individuals who were offered HCVST, 259 (35.8%) refused testing. The mean age of those who refused was 38.9 ± 12.3 years. Refusals were evenly distributed by gender, with 50.6% (131/259) male and 49.4% (128/259) female.

Through multivariable logistic regression analyses, factors associated with testing refusal were assessed. Participants aged 31–45 years had 1.9 times higher odds of refusal (95% CI: 1.32–2.73), and those aged 46–60 years had 2.14 times higher odds compared to those aged 18–30 years. Participants in the facility-based model had 1.87 times higher odds of refusal compared to those in the home-based delivery model.

In multivariable analysis, these findings remained consistent, with higher odds of refusal among participants aged 31–45 years (aOR = 2.00, 95% CI: 1.38–2.90) and 46–60 years (aOR = 2.18, 95% CI: 1.39–3.43), and among those in the facility-based model (aOR = 1.89, 95% CI: 1.37–2.60). Gender was not significantly associated with testing refusal. (Table 3)

**Table 3.**
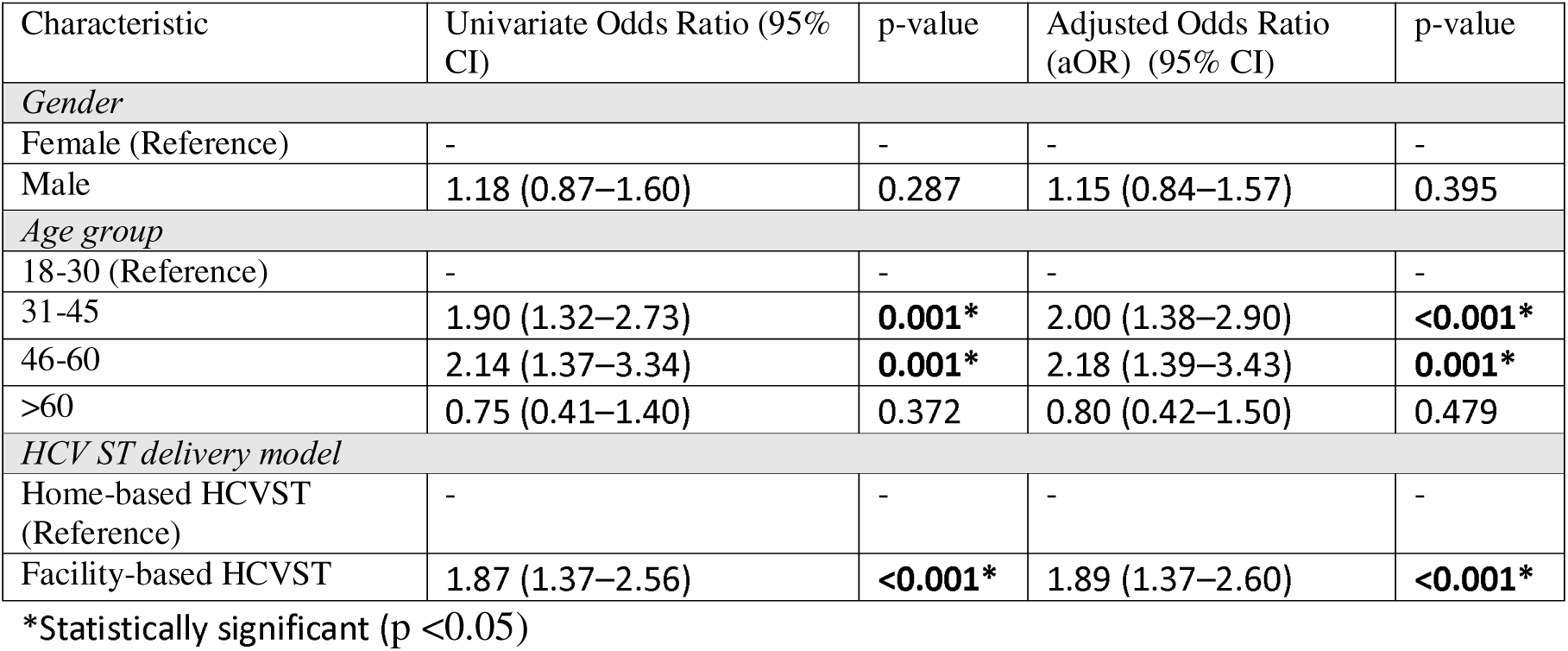
Factors associated with testing refusal; univariate and multivariable logistic regression analysis.

### Reasons for refusal

In addition to the factors associated with refusal above, we captured reported reasons for refusing to participate. The primary reason for refusal was a lack of time (29.3%, 76/259), with no significant difference between the facility-based model (28.9%, 48/166) and the home-based model (30.1%, 28/93; p = 0.84).. Fear of needle-pricks or finding a positive result (17.8%, 46/259) also showed no significant difference by site (19.3%, 32/166 at facility vs 15.1%, 14/93 in community; p=0.394). Statistically significant differences between the sites were observed for three reasons. The proportion of participants who preferred not to state a reason was significantly higher at facility (15.7%, 26/166) than the home-based site (6.5%, 6/93; p = 0.031). One in ten (10.0%, 26/259) felt no need for testing, though participants in the home-based model site were significantly more than at the facility (17.2%, 16/93 vs 6.0%, 10/166; p=0.004) and fasting (10.8%, 10/93 vs. 4.2%, 7/166; p = 0.042).

Other reported reasons included apreference to test elsewhere (8.1%, 21/259; p=0.229)and an inability to read the pamphlet instructions (7.7%, 20/259; p=0.338). A preference not to use the self-test was the least frequent reason (3.9%, 10/259) and did not differ significantly by site (p=0.976). (**Table 4**).

**Table 4.** Reasons of individuals who did not agree to participate.

| Reasons for refusing to participate | Total N=259 (%) | Facility-based site (Jwalapur 1) n = 166 (%) | Home-Based site (Jwalapur 2) n = 93 (%) | P-value (Chi-square/Fisher exact test) |
| --- | --- | --- | --- | --- |
| Do not have time to participate in the study | 76 (29.3) | 48 (28.9) | 28 (30.1) | 0.840 |
| Fearful (of finger prick or finding a positive result) | 46 (17.8) | 32 (19.3) | 14 (15.1) | 0.394 |
| Prefer not to say the reason | 32 (12.4) | 26 (15.7) | 6 (6.5) | 0.031* |
| Do not feel the need for testing | 26 (10.0) | 10 (6.0) | 16 (17.2) | 0.004* |
| Prefer to test elsewhere | 21 (8.1) | 16 (9.6) | 5 (5.4) | 0.229 |
| Could not read the pamphlet instructions | 20 (7.7) | 11 (6.6) | 9 (9.8) | 0.338 |
| Fasting | 17 (6.6) | 7 (4.2) | 10 (10.8) | 0.042* |
| Other<sup>#</sup> | 11 (4.2) | 9 (5.4) | 2 (2.2) | 0.355 |
| Prefer not to use the self-test | 10 (3.9) | 7 (4.2) | 3 (3.2) | 0.976 |
| #Includes: Already tested in another hospital, recent pregnancy, hesitant to give blood sample, etc) |  |  |  |  |
| * Statistically significant (p <0.05) |  |  |  |  |

Qualitative feedback indicated that “lack of time” was driven by competing health priorities and misconceptions. Facility-based participants prioritized their primary ailments, perceiving opportunistic screening as a significant additional time commitment. Furthermore, many overestimated the complexity and duration of the test, unaware that the process requires only 15–20 minutes. ‘Fearfulness’ was a multi-faceted barrier, encompassing both the immediate fear of the finger-prick and the psychological fear of receiving a positive result and the perceived stigma associated with a diagnosis.

### HCVST case detection proportion

Among participants in the HCV self-testing (HCVST) sites, the combined HCV antibody positivity rate was 1.1% (5/465; 95% CI: 0.35%–2.49%). The facility-based HCVST was slightly higher (1.8%, 4/227; 95% CI: 0.50%–4.45%) compared to the home-based HCVST site (0.4%, 1/238; 95% CI: 0.01%–2.32%). In contrast, no HCV antibody–positive cases were identified in the SoC site (0/11). All five HCVST reactive individuals were successfully linked to confirmatory testing and clinical evaluation. Laboratory-based anti-HCV ELISA and HCV RNA testing were performed for all five individuals.Three had detectable HCV RNA, confirming active HCV infection, and were initiated on direct-acting antiviral (DAA) therapy. All three completed treatment and achieved sustained virological response, corresponding to a 100% cure rate among those treated.

## Discussion

HCVST delivered through facility and home-based models was associated with a substantially higher number of tests conducted compared with routine programme delivery (SoC), where only 11 tests were delivered over the same time period. The home-based HCVST distribution model had slightly lower OPD-footfall-standardized operational reach, but higher uptake among those offered testing than the facility-based model. Despite these differences in engagement however, neither model identify a substantial HCV case burden, reflected in the overall low case detection proportion of HCV antibody positivity, with only five reactive cases identified across both intervention sites. This difference should not be attributed solely to self-testing versus conventional testing. Each delivery model was implemented in a single site, and HCVST was actively offered through study-supported facility or household delivery, whereas testing at the SoC site depended on routine programme workflows. Differences in implementation intensity and site-level factors may therefore have contributed to the low number of tests recorded at the SoC site. These results align with established evidence that self-testing, integrated across diverse distribution models, consistently outperforms routine testing. In HIVST, facility-based models in Malawi showed 8.5 times higher odds of testing, while community-based campaigns achieved up to 84% coverage (19,20). Linkage to confirmatory testing and treatment initiation also remains robust, particularly when using assisted HIVST (21). Evidence specific to HCVST further supports these gains; Wang et al. reported substantially higher testing among MSM in China compared to SoC (22), while a multi-country RCT in Georgia, Malaysia, and Pakistan demonstrated higher uptake and linkage to care in HCVST groups versus controls (23).

Collectively, these studies suggest that integrating self-testing into diverse delivery models is a robust approach to overcoming the limitations of conventional diagnostic pathways.

The differences in uptake observed between the two HCVST models likely reflect the mode of delivery, contextual factors and populations reached. While more individuals were offered HCVST at the facility-based site, which may reflect the relative ease of approaching individuals within facility-based settings, the home-based model achieved higher uptake, which may reflect the convenience of in-home testing and support from ASHAs. This reaffirms that accessible door to door household service delivery mechanisms through a trusted community peer can engage populations (10).

Participants in the facility-based model had nearly double the odds of refusing testing, compared with the home-based model; however, because each delivery model was implemented at a single site, this difference cannot be attributed to the delivery model alone. Additionally, participants over age 30 were twice as likely to refuse testing compared to those under 30. These findings suggest that a door-to-door delivery model may warrant further evaluation as an approach to improve testing acceptance, while tailored strategies may be needed to reach older populations of more than 30 years.

Our results align with recent evidence from Vietnam, where community-based distribution of HCVST effectively identified the most people with HCV and reached a higher proportion (83.8%) of first-time testers than facility-based care (24). As a “self-care” option, HCVST bypasses the need for formal healthcare interactions, expanding reach to those who might otherwise avoid seeking existing services at the facility. (22,24). Conversely, research in Malawi and China indicates that routine facility-based testing models often act as bottlenecks due to “institutional barriers”, facilities are often underutilized by men and high-risk groups due to stigma of being seen at specialised clinic, along long wait times and the opportunity cost of missing work (25,26). By removing testing from the “sick-person” context, door-to-door or peer-led models “normalize” screening and can increase testing uptake by 5.8% to 37% compared to facility-based approaches (27).

This study did not collect economic data and therefore cannot determine whether facility-based or home-based HCVST was cost-effective. Previous economic evaluations and modelling studies suggest that HCVST may be economically attractive under some conditions, but its value is likely to depend on background HCV prevalence, test-kit costs, staff time, implementation intensity, confirmatory-testing requirements, and linkage to treatment (28, 29). Given the low number of infections detected in this study, universal or untargeted HCVST cannot be recommended on the basis of these findings alone. Future evaluations should assess cost per person tested, cost per active infection diagnosed, cost per person treated and cured, and the budget impact of integrating HCVST into routine services.

In our context, the higher uptake of the home-based model site suggests the potential for ASHAs to support HCVST delivery through existing community health infrastructure. Community health worker involvement may offer a feasible approach to improving accessibility; however, the present study cannot isolate the effect of ASHA involvement from other site- and implementation-level factors. Integration into routine services would therefore require further evaluation of training requirements, supervision, commodity supply, compensation, and the opportunity costs of adding HCVST to existing responsibilities (30–33).

However, the overall low case detection proportion, and that a third of participants had no reported risk factors, suggests that these interventions may not have been sufficiently targeted at risk groups within urban slum; general population provision of screening was not sufficient to reach high-risk groups. Although it is important to note that there is no baseline epidemiological data specific to these communities. This underscores the need for reliable estimates of disease burden before scaling up HCVST. Prior research has shown that self-testing is most impactful in high-prevalence groups, such as PWIDs (34,35).

### Limitations

A key limitation was eligibility criteria of needing to reside in the urban slum for a minimum of 6 months prior to study inclusion. Although sparse, risk factors associated with slum living include occupation risk exposures of recent rural-urban migrants; excluding these populations may have been partially responsible for the low case detection proportion within this study.

Conversely, restricting study eligibility to adults may have led to an underestimation of the OPD-footfall-standardized operational reach, as the OPD footfall denominator included individuals younger than 18 years. Community-level reach could not be directly estimated because unique community contacts were not prospectively recorded. OPD footfall was therefore used as a proxy for service volume and should not be interpreted as the proportion of the eligible community population reached. The reliance on self-reported risk behaviors may also have introduced social desirability bias; for example, no participant reported drug use. As initial pilots, there was no data collected on the adoption, long-term implementation, or maintenance collected. The low positivity rate restricted the study’s ability to assess linkage to care, treatment initiation, and outcomes. Future research is needed to evaluate the long-term sustainability of these models and the institutional capacity of primary healthcare centers to integrate them into routine practice.

## Conclusion

HCV self-testing was associated with improved testing and uptake when delivered through both facility- and home-based models compared with routine programme delivery, although differences between sites may also reflect variation in implementation intensity and local context. Trusted community peer (ASHA)-led door-to-door distribution showed higher testing uptake, although this finding may also reflect site-level differences. Decentralizing HCV services by integrating HCVST into primary healthcare and task-shifting to community health workers may strengthen national diagnostic programming (30,36). However, given the low case detection proportion in this study, future work is required to 1) better understand HCV burden in urban slums and 2) apprioriately target HCVST strategies to guide rational allocation of HCVST kits, ensuring that testing reaches those most in need.

## Data Availability

All data produced are available as supporting documents with this manuscript.

## Supplements

**S1. Baseline data assessment at sites randomly selected for study inclusion.**

**S2. The TIDieR (Template for Intervention Description and Replication) Checklist.**

**S3. Data file supporting analysis.**

