## Supplement 2 for "Reach and uptake of facility- and home-based hepatitis C self-testing in urban slums in Haridwar, India; A Comparative Implementation Study"

| **Supplementary material: S2 Intervention Implementation details based on TIDieR checklist** | | | |
| --- | --- | --- | --- |
| **Intervention Characteristics** **†** | **Intervention site 1** | **Intervention site 2** | **Control site** |
| **Name of the intervention** | Facility-based HCV self-testing | Community-based HCV self-testing | Standard of care (SOC) |
| **Why:**  Rationale for the intervention (Theory of Change) | Haridwar is a rapidly urbanizing district of Uttarakhand, with a significant slum population (25%) and history of high HCV prevalence due to unsafe injection practices. Urban Slums often house migrants who are vulnerable due to systemic risk behaviours and structural barriers. The provision of HCVST kits through two delivery models along with SOC (health education and awareness, free diagnostic services (RDTs) and referral) to increase testing uptake among vulnerable populations compared to the SOC under NVHCP of India. Anticipated outcomes were determining and comparing the reach and effectiveness of the two models against each other and against the SOC, case finding of HCV cases, linking to higher treatment centres and follow up with Sustained Virologic Response (SVR) to ensure treatment completion and viral clearance. | | |
| **What:**  *Materials* | Across both intervention sites, IEC material: Posters and pamphlets with information on Hepatitis C virus and disease signs and symptoms and treatment along with Instructions for Use (IFU) with pictorial representation of steps to use the self-testing kits in Hindi language were distributed at PHCs. An example of the pamphlet is included below.  HCVST kits- The First Response HCV Card Test (Premier Medical Corporation Pvt Ltd., Gujarat, India) was used for self-testing, with MONOLISA HCV Ag-Ab ULTRA V2 as the reference test.  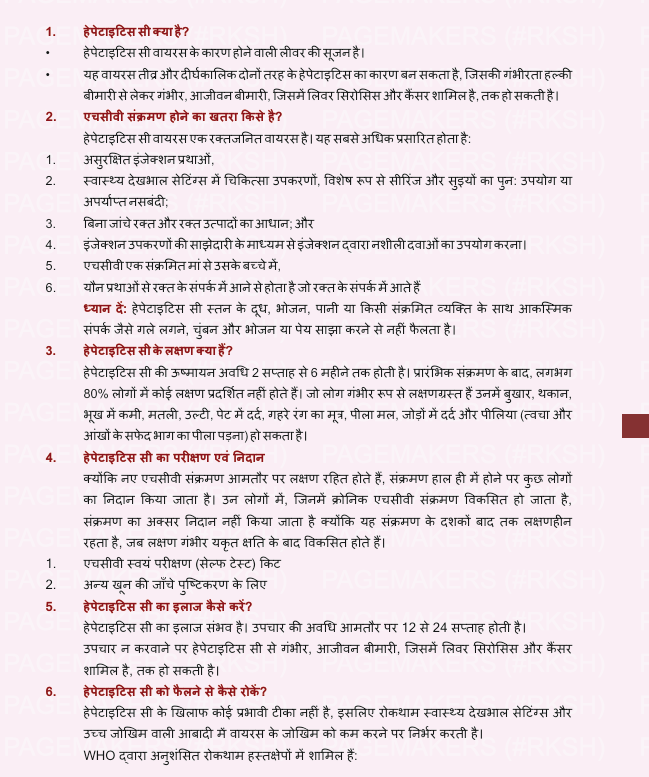  Example of print media (Pamphlet) for demand creation created by study team. | | IEC materials at UPHC Kankhal as provided under NVHCP. |
| **What**:  *Procedures* | 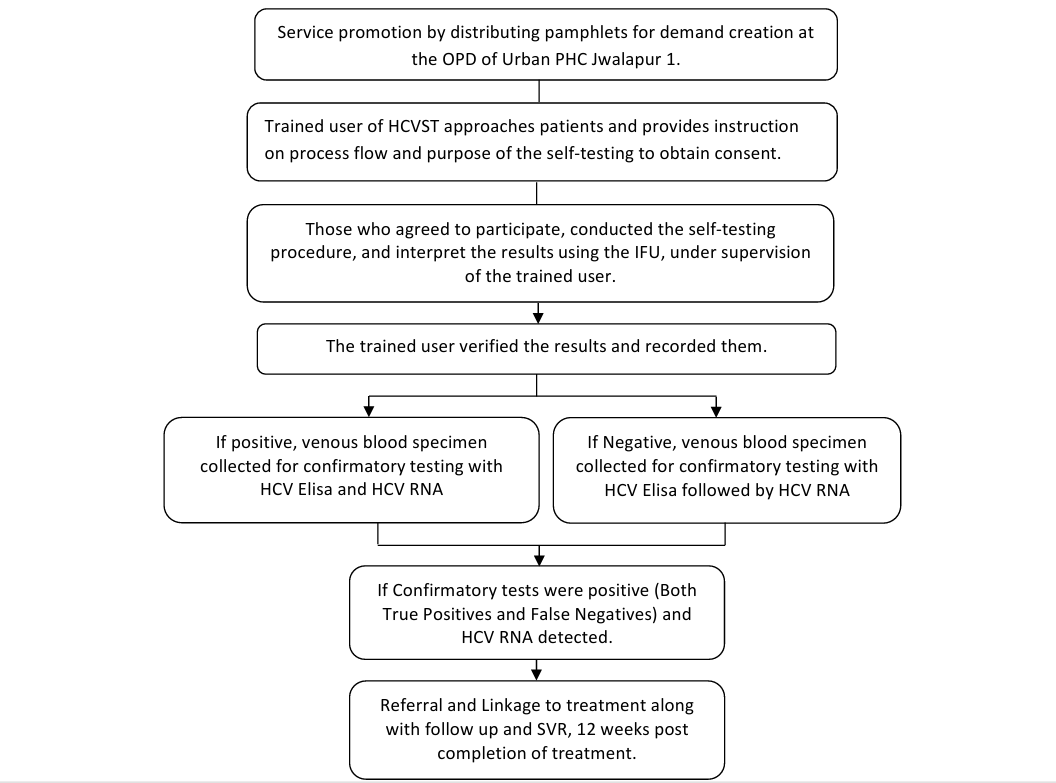 | 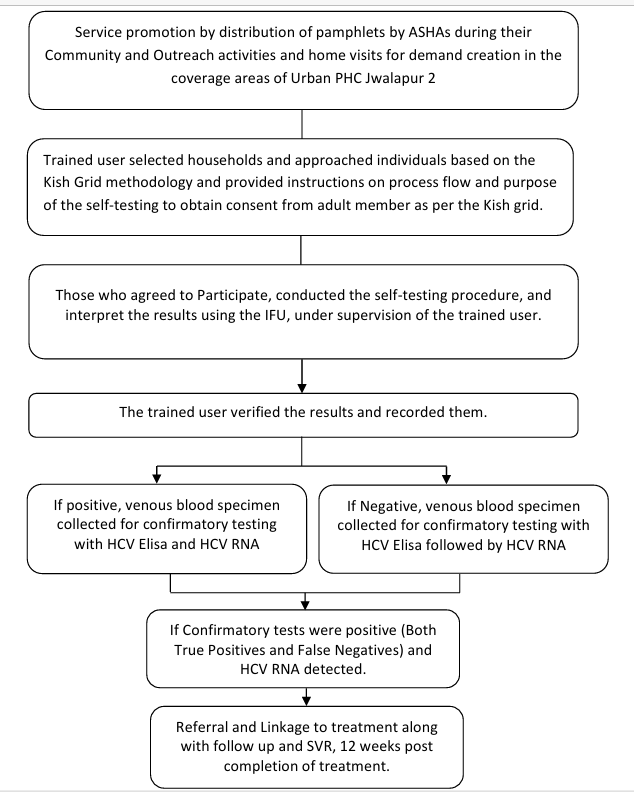 | - Routine NVHCP activities Awareness Generation and Behaviour change communication - Free screening, diagnosis, and treatment for both hepatitis B and C (especially, screening of pregnant women) - Provision of linkages, including with private sector and not for profit institutions, for diagnosis and treatment. |
| **Who** **provided:** | Trained User of HCVST and Lab Technician with assistance from medical officer (MO) at the Urban PHC at Jwalapur 1 | Trained User of HCVST and Lab Technician along with the help of Accredited Social Health Activists (ASHAs) of Urban PHC Jwalapur 2 area. | Community Health Officer (CHOs), Auxiliary Nursing Midwives (ANMs) and ASHAs |
| **How:**  *Mode of delivery; individual or group:* | Demand creation and service promotion happened one on one, with individuals (patients) visiting the OPD. Consecutive sampling method was used for recruitment of participants  Follow-up was through telephone. | Demand creation and service promotion happened one on one during the home visits in the community by the team. Households and their members were selected based on the Kish Grid methodology. | Routine OPD services. |
| **Where:** | At the Selected facility of Urban PHC Jwalapur 1, Haridwar | Households of area covered by UPHC Jwalapur 2, Haridwar | Urban PHC Kankhal, Haridwar |
| **When and how much:** | The service was available during the OPD hours from 9 am to 2 pm, Monday to Saturday.  One test kit for one participant. | The households were visited on weekdays between 11 am to 6 pm. On weekends, 12 noon to 5 pm. | During the OPD hours at the facility at Kankhal |
| **Tailoring:** | Except for the site of approach and delivery of the service, there was no difference in the intervention | | HCV self-testing kits were not part of the intervention. |
| **Modifications:** | In response to the observed reluctance among the target population, the questionnaire aimed at capturing reasons for self-testing refusal was truncated and contextually adapted to ensure greater acceptability. | |  |

**†** Not represented in the table is how well (planned and actual) the intervention was adhered to (fidelity), because there was no formal assessment of this.

Procedure flow for community based self-testing

Service promotion by distribution of pamphlets by ASHAs during their Community and Outreach activities and home visits for demand creation in the coverage areas of Urban PHC Jwalapur 2

The trained user verified the results and recorded them.

Referral and Linkage to treatment along with follow up and SVR, 12 weeks post completion of treatment.

If Confirmatory tests were positive (Both True Positives and False Negatives) and HCV RNA detected.

If Negative, venous blood specimen collected for confirmatory testing with HCV Elisa followed by HCV RNA

If positive, venous blood specimen collected for confirmatory testing with HCV Elisa and HCV RNA

Trained user selected households and approached individuals based on the Kish Grid methodology and provided instructions on process flow and purpose of the self-testing to obtain consent from adult member as per the Kish grid.

Those who agreed to Participate, conducted the self-testing procedure, and interpret the results using the IFU, under supervision of the trained user.

Procedure flow for facility based self-testing

Referral and Linkage to treatment along with follow up and SVR, 12 weeks post completion of treatment.

If Confirmatory tests were positive (Both True Positives and False Negatives) and HCV RNA detected.

If Negative, venous blood specimen collected for confirmatory testing with HCV Elisa followed by HCV RNA

The trained user verified the results and recorded them.

Service promotion by distributing pamphlets for demand creation at the OPD of Urban PHC Jwalapur 1.

Those who agreed to participate, conducted the self-testing procedure, and interpret the results using the IFU, under supervision of the trained user.

Trained user of HCVST approaches patients and provides instruction on process flow and purpose of the self-testing to obtain consent.

If positive, venous blood specimen collected for confirmatory testing with HCV Elisa and HCV RNA
