## Supplement 1 for "Reach and uptake of facility- and home-based hepatitis C self-testing in urban slums in Haridwar, India; A Comparative Implementation Study"

**S1. Baseline data assessment at sites randomly selected for study inclusion.**

The baseline data from July 2023 to December 2023, regarding human resources, trainings, patient footfalls, population coverage, availability and utilization of hepatitis C screening kits, diagnostics, and directly acting antivirals were comparable across all three Primary Health Centres (**Table S1)**.

**Table S1: Baseline data of three PHCs for Hepatitis C prevention, diagnosis, and management under NVHCP**

| **NVHCP activities** | **Jwalapur 1 (Facility-based site)** | **Jwalapur 2 (Community-based site)** | **Kankhal (Control site)** |
| --- | --- | --- | --- |
| Awareness generation and BCC | Not done | Not done | Not done |
| Immunization for Hep B- BD, high risk groups, health care workers | Available | Available | Available |
| Injection safety- re-use prevention syringes | Available | Available | Available |
| Diagnosis tests: serological tests | Available (only for ANC and symptomatic patients with jaundice and deranged LFT reports) | Yes (only for ANC patients) | Yes (only for ANC patients) |
| Confirmation: molecular tests | Available | Available | Available |
| Treatment of uncomplicated and Complicated cases | Not available  (Referred to CHC Jwalapur) | Not available  (Referred to CHC Jwalapur, JD hospital) | Not available  (Referred to CHC Jwalapur, JD hospital) |
| Availability of DAA drugs | Absent | Absent | Absent |
| Referral and linkages | Present | Present | Present |
| Training and capacity building of health care staff | Not done | Not done | Not done |
| Hepatitis information and Management portal | Not used | Not used | Not used |
| Surveillance of viral hepatitis | Not done | Not done | Not done |
| Monitoring and Evaluation | Not done | Note done | Note done |

The table displays the current state of hepatitis C prevention, diagnosis, and management under the NVHCP. Serological tests for hepatitis C screening and diagnosis are only conducted for antenatal pregnant women and symptomatic patients. Services such as awareness generation and behaviour change communication are practically non-existent. There is also a lack of availability of Direct Acting Antiviral (DAA) drugs for treatment. Any positive cases are referred to the nearest Community Health Centre in Jwalapur for further treatment.
